# A longitudinal analysis of incidence of hypertension and blood pressure measurements by age of migration among older Hispanic men and women

**DOI:** 10.1101/2024.04.03.24305302

**Authors:** Brandon S. Walker, Norman J Waitzman, Evan V. Goldstein, Megan E. Vanneman, Alan Taylor Kelley, Fernando A. Wilson

**Affiliations:** Department of Population Health Sciences, University of Utah, Salt Lake City, Utah; Department of Economics, University of Utah, Salt Lake City, Utah; Division of General Internal Medicine, University of Utah, Salt Lake City, Utah

**Keywords:** Aging, Blood Pressure, Hispanic or Latino, Hypertension, Longitudinal Studies, Social Class, United States

## Abstract

**Objective:** To conduct a longitudinal analysis of incidence of self-reported hypertension and blood pressure measurements among foreign-born Hispanics by age of migration, compared to US-born populations.

**Methods:** The sample was drawn from 2002-2018 of the Health and Retirement Study and included 22,909 individuals. Subsets of this sample were used to conduct a longitudinal analysis of the incidence of hypertension and mean blood pressure measurements.

**Results:** Foreign-born Hispanic women migrating at age 40 and older had a greater incidence of hypertension and a greater increase in systolic blood pressure as they aged compared to US-born Whites.

**Discussion:** In contrast to the Hispanic Paradox that suggests better health among Hispanic immigrants despite lower socioeconomic status, this was not observed among older Hispanic immigrants for hypertension. Furthermore, older Hispanic women who migrated later in life had a greater incidence of hypertension and greater increases in systolic blood pressure as they aged compared to US-born White women.

## Introduction

Hispanic immigrants in the United States often have better health outcomes and longer life expectancies compared to non-Hispanic Whites, despite their lower socioeconomic status. ^1,2^ This paradox of better health, often called the “Hispanic paradox” has been attributed to several factors including healthier lifestyles, greater levels of informal social support, and a selection process where healthier individuals are more likely to migrate to the United States for employment opportunities. ^3–6^ However, these health advantages generally decline with increased years of residency in the United States as immigrants adopt less healthy lifestyles and are exposed to stressors related to acculturation. ^6,7^ Immigrant health is also shaped by the age of migration to the United States. While those who migrate earlier in life are motivated by employment opportunities, those migrating later in life are more likely to be motivated by family reunification and, therefore may be less health-selected. ^8,9^ Additionally, those who migrate late in life may have more difficulty adapting to their new environments compared to those that migrated earlier. ^10,11^ These difficulties may include loss of social networks and limited English proficiency, resulting in greater social isolation. ^4,12–14^ Migration later in life has been associated with a greater risk of functional disability, ^15,16^ depression, ^17–21^, cognitive decline, ^22–24^, and poorer self-rated health^25^, although the mechanisms are not clearly understood.

Consistent with the Hispanic Paradox, Hispanic immigrants have lower rates of hypertension compared to US-born non-Hispanic Whites. ^5,26,27^ These risks of hypertension generally increase among Hispanic immigrants with longer residency in the United States. ^6,28,29^ However, these health advantages for hypertension are not observed among older Hispanic immigrants who have a greater risk of hypertension. ^30,31^ Walker and Waitzman found that those with fewer years of residency had the lowest risk of hypertension compared to non-Hispanic Whites. ^31^ However, among foreign-born Hispanics aged 65 years and older, those with less than 10 years of residency had a significantly greater risk of hypertension, while those with greater than 10 years of residency had risks similar to US-born non-Hispanic Whites. ^31^ This counterintuitive finding of greater hypertensive risk among those with the fewest years of residency among older foreign-born Hispanics suggests risk factors associated with migration later in life. However, prior research is limited by the cross-sectional design of the data used and, thus, it cannot be determined if these findings reflect differences in the age cohorts rather than the trajectory of individuals as they age. It is currently unknown how age of migration influences the risk of hypertension among older Hispanic immigrants. It is also unknown what mechanisms might be responsible for differences in risk by age of migration. To address this gap in knowledge, our study uses nationally representative, longitudinal data to provide new insights into the role of age of migration in hypertensive risk as well as possible mechanisms that act through age of migration to influence the risk of hypertension. We utilize data on self-reported hypertension and blood pressure measurements to compare the risk of hypertension of foreign-born Hispanics, by age of migration, to US-born populations.

## Methods

### Study design

Study design: Data were drawn from years 2002-2018 of the Health and Retirement Study (HRS). The HRS is a longitudinal dataset that is publicly available and nationally representative of non-institutionalized adults ages 50 and older in the United States. The years 2002-2018 were chosen because of the availability of household income represented as a percentage of federal poverty level. The sample included foreign and US-born Hispanics as well as US-born non-Hispanic Whites (henceforth called US-born Whites). Applying these criteria identified 24,196 individuals of which 22,909 had no missing data at baseline (first year of observation). Race/ethnicity was self-reported. Hispanics were identified in the HRS with an affirmative response to the question “Do you consider yourself Hispanic or Latino”.

Measurements: Dependent variables are self-reported previous diagnosis of hypertension as well as diastolic and systolic blood pressure measurements. Participants in the HRS alternate between a face-to-face and telephone interview every other HRS wave. Since the HRS collects information bi-annually, a face-to-face interview occurs every four years. Hypertension was defined as having a systolic blood pressure of at least 140 mm Hg or a diastolic blood pressure of at least 90 mm Hg or the reporting current usage of antihypertensive medication based on guidelines of the American Heart Association (AHA). ^32^ Blood pressure measurements were taken by trained field interviewers at the time of the face-to-face interview. Three blood pressure measurements were taken, the second and the third readings were then averaged for the blood pressure measurement based on recommendations from the AHA. ^33,34^ Interviews were conducted in English or Spanish.

The exposure groups used in the analysis included: foreign-born Hispanics: with age of migration groups of 0-19, 20-39, 40+ years, US-born Whites, and US-born Hispanics. The analysis was stratified by sex due to differences in hypertension between men and women. ^35^ Socioeconomic status measured in income and educational attainment were included in the analysis because both lower educational attainment and income are associated with hypertension. ^36^ Educational attainment was categorized as less than high school, high school, and greater than high school. Income was categorized into three categories: 0-149% federal poverty level (FPL) in order to capture poverty and near poverty income while 150-299% FPL and 300%+ FPL were used to capture the remaining distribution of income. Health insurance was included in the model because of its association with undiagnosed hypertension which impacts self-reported hypertension status and because of its association with lower blood pressure measurements. ^37,38^ Lifestyle and behavioral risk factors of smoking status, alcohol use, depression, and Body Mass Index (BMI) are associated with greater risk for hypertension. ^39–42^ Current smoking and alcohol use were included as binary variables. Depression was defined as a binary variable using the 8-item short version of the Center for Epidemiologic Studies-Depression Scale collected in the HRS. Participants with a score of 3 or greater were categorized as at risk for clinical depression. ^43^ BMI was defined as a categorical variable and categorized into the following groups based upon measured height and weight: underweight/normal weight (0-24.9), overweight (25.0-29.9) and obese (≥ 30.0). Blood pressure medication was included in the analysis of blood pressure measurements since it reduces blood pressure.

Statistical analysis: All analysis was conducted using Stata 17.0. Summary statistics are provided for the 22,909 participants at baseline (first year of observation). Subsamples of these individuals were examined with Cox regression and mixed effects linear regression. The analysis was appropriately specified using the HRS provided weights, strata, and probability sampling units to account for the complex sample design. Sample weights were adjusted for missing values using inverse probability weights for both Cox and mixed effects regression.

Cox regression was used to examine the incidence of self-reported hypertension while controlling for covariates at baseline levels. Baseline covariates were used to prevent violation of the proportional hazards assumption used in Cox regression. Although methods exist to adjust for time varying covariates in Cox regression, these methods are not supported with use of complex survey data. Age was used as the time scale in the Cox regression analysis and therefore not included as a covariate in the model. ^44,45^ The proportional hazards assumption was examined using a global test of Schoenfeld residuals. The model fit was assessed by plotting the Cox-Snell residuals. Individuals with hypertension at baseline were excluded from the analysis. Excluding individuals with hypertension at baseline resulted in the inclusion of 12,062 individuals and a total sample size of 46,637. Both death and lost to follow up were treated as censoring events. Because Cox regression treats censoring events as uninformative (not related to the outcome being examined) an analysis of mortality and a competing risk regression analysis were also conducted. Cox regressions were used to examine mortality to determine how survivorship bias may have impacted the results. Competing risk regression was also used, however because adjustment of standard errors due to the complex survey design is not supported for competing risk regressions, these results should be interpreted cautiously since standard errors may be underestimated. Mortality in the HRS was determined through the National Death Index.

Mixed effects regression analysis was conducted on blood pressure measurements. The analysis included all individuals with measurements regardless of hypertension status. The HRS currently provides blood pressure measurements for years 2006-2018. Mixed effects analysis included 16,690 individuals for a sample size of 35,947 observations. Age was centered at 50 years for the mixed effects regression to allow for a more meaningful interpretation of regression results whereby race/nativity coefficients (without interactions) can be interpreted as the difference in predicted values in comparison to US-born Whites at age 50. An age squared term was also included in the regression formula to allow for a non-linear relationship between age and blood pressure measurements. A cross level interaction between age variables and race/nativity groups were used to capture group differences in trajectory over time. The group interaction with age represents the slope at age 50 while the interaction with age squared represents the curvature over time. Model fit was assessed with a likelihood ratio test. The mixed effects model included a random intercept and assumed an unstructured covariance matrix.

For both regressions, variables were added in a stepwise fashion to determine the influence of socio-economic, health insurance, and behavioral factors on group differences. Risk factors were not included in the first model (Model 1). In Model 2, educational attainment and household income were added to the analysis. In Model 3, health insurance status was added. Model 3 also included use of blood pressure medication in the analysis of blood pressure measurements. Finally, in Model 4, behavioral risk factors of smoking, alcohol use, BMI, and depression were added to the analysis.

## Results

### Summary statistics

Overall, the average age of participants at baseline was statistically lower among US Hispanics and foreign-born Hispanics who migrated before age 40 compared to US-born Whites (Table 1). The distribution of men and women was statistically similar between race/nativity groups except for foreign-born Hispanics that migrated ages 20-39 who had a greater proportion of men compared to US-born Whites.

**Table 1.**
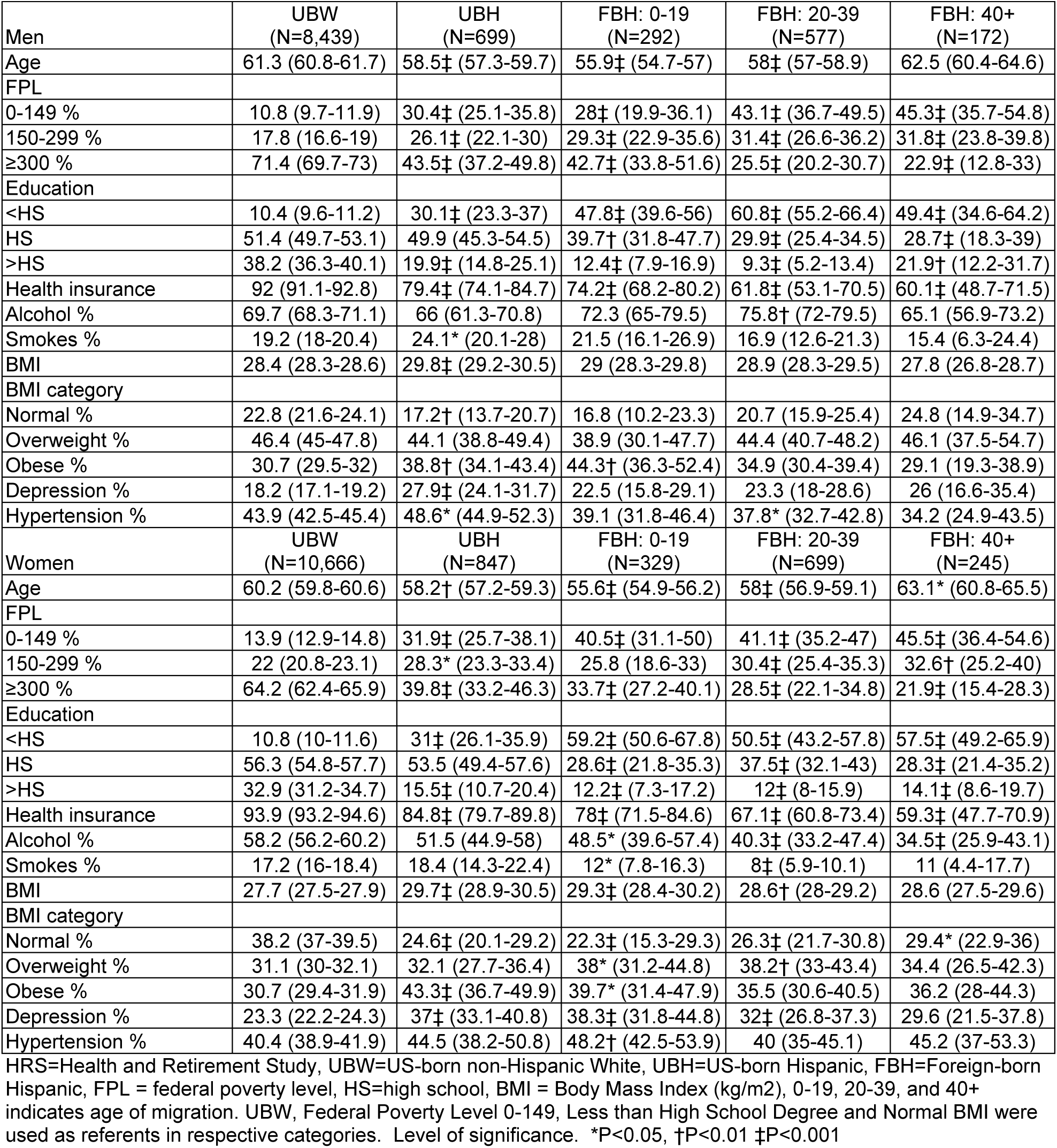
Summary statistics of included individuals as baseline stratified by sex. HRS 2006-2018.

All Hispanic groups had lower socioeconomic status measured in income as a percentage of federal poverty level and educational attainment. The proportion of participants with household incomes within 0-149% federal poverty level (FPL) and educational attainment less than high school was statistically higher (p values<0.001) among all Hispanics compared to US-born Whites, with greatest levels among foreign-born Hispanics. Similarly, levels of health insurance were lower among US and foreign-born Hispanics compared to US-born Whites, with the lowest levels of health insurance among foreign-born Hispanics migrating after age 40.

Behavioral and lifestyle measurements differed across race/nativity groups (Table 1). The number of those currently smoking was generally similar among race/nativity groups except for US-born Hispanic men who had a greater risk of smoking compared to US-born White men, and foreign-born Hispanic women who migrated ages of 20-39 who had a lower risk compared to US-born White women. Alcohol use was higher among foreign-born Hispanic men who migrated between the ages of 0-19 compared to US-born White men but similar for other race/nativity groups. Alcohol use was lower for all foreign-born Hispanic women compared to US-born White women. The level of obesity was greater among US-born Hispanic and foreign-born Hispanic men migrating between ages 0-19 compared to US-born White men. Among Women, obesity was greater for foreign-born Hispanic women migrating between ages 0-19 and 20-39 compared to US-born White women. US-born Hispanic men had a greater proportion of those with depression compared to US-born White men. US-born Hispanic and foreign-born Hispanic women with ages of migration 0-19, and 20-39 had greater rates of depression compared to US-born White women.

Prevalence of hypertension at baseline varied among race/nativity groups. US-born Hispanic men had a greater prevalence of self-reported hypertension at 48.7% (95% CI=45.0, 52.4, p=0.018) while foreign-born Hispanic men migrating between ages 20-39 had a lower prevalence at 37.8% (95% CI=32.8, 42.9, p=0.029) compared to US-born White men at 44% (95% CI=42.5, 45.5). Foreign-born Hispanic women migrating between ages 0-19 had a greater prevalence of hypertension at 48.3% (95% CI=42.6, 53.9, p=0.010) compared to US-born White women at 40.6% (95% CI=39.1, 42.1).

### Cox regression of hypertension incidence

Cox regression analysis was conducted on individuals without self-reported hypertension at baseline. Hypertension incidence was greater among US-born and foreign-born Hispanic men that migrated ages 0-19 with hazard ratios (HRs) of 1.57 (95% CI=1.22, 2.02, p=0.001) and 1.43 (95% CI=1.16, 1.76, p=0.001) compared to US-born White men in the unadjusted model (Table 2, Model 1). This greater risk relative to US-born Whites persisted in US-born Hispanic men even after adjusting for baseline socioeconomic, health insurance, and behavioral risk factors (Model 4) with a HR of 1.36 (95% CI=1.07, 1.73, p=0.014), however, the HR was no longer statistically greater for foreign-born Hispanic men that migrated ages 0-19 compared to US-born White men after adjusting for socioeconomic status (Model 2). Among women, both US-born and foreign-born Hispanics migrating at age ≥ 40 years had a greater risk of developing hypertension compared to US-born White women with HRs of 1.56 (95% CI=1.34, 1.81, p<0.001) and 1.68 (95% CI=1.26, 2.25, p=0.001) in the unadjusted model respectively. However, after adjusting for education and income, foreign-born Hispanic women migrating at age ≥ 40 were no longer statistically different than US-born White women, in contrast, US-born Hispanic women maintained this greater risk even when adjusting for baseline socioeconomic status, health insurance, and behavioral factors with a HR of 1.2 (95% CI=1.01-1.43, p=0.043).

**Table 2.** Cox regression results in incidence of self-reported hypertension. Results reported in hazard ratios. HRS 2002-2018.

|  | Model 1 | Model 2 | Model 3 | Model 4 |
| --- | --- | --- | --- | --- |
| **Men (N=19,741)** |  |  |  |  |
| UBH | 1.57† (1.22, 2.02) | 1.43† (1.12, 1.84) | 1.39† (1.09, 1.77) | 1.36* (1.06, 1.73) |
| FBH: 0-19 | 1.33 (0.99, 1.80) | 1.16 (0.87, 1.54) | 1.11 (0.84, 1.48) | 1.14 (0.86, 1.50) |
| FBH: 20-39 | 1.43† (1.16, 1.76) | 1.19 (0.95, 1.50) | 1.13 (0.89, 1.43) | 1.07 (0.84, 1.38) |
| FBH: ≥ 40 | 1.15 (0.82, 1.62) | 1.01 (0.69, 1.49) | 0.94 (0.63, 1.42) | 0.89 (0.59, 1.35) |
| Federal poverty level |  |  |  |  |
| 150 to 299 % |  | 0.89 (0.71, 1.10) | 0.91 (0.73, 1.13) | 0.91 (0.74, 1.12) |
| 300+ % |  | 0.87 (0.73, 1.03) | 0.91 (0.77, 1.08) | 0.89 (0.76, 1.05) |
| Education |  |  |  |  |
| High school |  | 0.95 (0.81, 1.11) | 0.97 (0.82, 1.15) | 1.01 (0.86, 1.19) |
| More than high school |  | 0.71† (0.58, 0.87) | 0.74† (0.60, 0.91) | 0.81* (0.66, 1.00) |
| Health insurance |  |  | 0.76* (0.62, 0.94) | 0.76* (0.61, 0.93) |
| Alcohol use |  |  |  | 1.07 (0.93, 1.22) |
| Smoking |  |  |  | 1.17* (1.01, 1.36) |
| BMI category |  |  |  |  |
| Overweight |  |  |  | 1.36‡ (1.21, 1.52) |
| Obese |  |  |  | 1.99‡ (1.73, 2.30) |
| Depression |  |  |  | 1.05 (0.90, 1.22) |
| **Women (N= 26,896)** |  |  |  |  |
| UBH | 1.56‡ (1.34, 1.81) | 1.26* (1.06, 1.50) | 1.26† (1.06, 1.50) | 1.20* (1.01, 1.43) |
| FBH: 0-19 | 1.29 (0.87, 1.91) | 0.98 (0.68, 1.41) | 0.98 (0.68, 1.42) | 0.92 (0.62, 1.36) |
| FBH: 20-39 | 1.22 (0.93, 1.61) | 0.94 (0.73, 1.20) | 0.94 (0.74, 1.20) | 0.92 (0.72, 1.19) |
| FBH: ≥ 40 | 1.68† (1.25, 2.25) | 1.27 (0.97, 1.68) | 1.28 (0.96, 1.72) | 1.28 (0.94, 1.73) |
| Federal poverty level |  |  |  |  |
| 150 to 299 % |  | 0.88* (0.78, 0.99) | 0.88* (0.78, 0.99) | 0.88* (0.78, 0.98) |
| 300+ % |  | 0.74‡ (0.64, 0.84) | 0.73‡ (0.64, 0.84) | 0.77† (0.66, 0.89) |
| Education |  |  |  |  |
| High school |  | 0.73‡ (0.64, 0.84) | 0.73‡ (0.64, 0.84) | 0.74‡ (0.65, 0.85) |
| More than high school |  | 0.66‡ (0.57, 0.77) | 0.66‡ (0.57, 0.77) | 0.70‡ (0.60, 0.82) |
| Health insurance |  |  | 1.03 (0.83, 1.28) | 1.06 (0.85, 1.32) |
| Alcohol use |  |  |  | 0.94 (0.84, 1.05) |
| Smoking |  |  |  | 1.06 (0.93, 1.19) |
| BMI category |  |  |  |  |
| Overweight |  |  |  | 1.43‡ (1.29, 1.59) |
| Obese |  |  |  | 1.83‡ (1.63, 2.06) |
| Depression |  |  |  | 1.00 (0.89, 1.13) |

HRS=Health and Retirement Study, UBW=US-born non-Hispanic White, UBH=US-born Hispanic, FBH=Foreign-born Hispanic, FPL = federal poverty level, HS=high school, BMI = Body Mass Index (kg/m<sup>2</sup>), 0-19, 20-39, and ≥ 40 indicates age of migration. UBW, Federal Poverty Level 0-149, Less than High School Degree and Normal BMI were used as referents in respective categories. Level of significance. \*P<0.05, †P<0.01 ‡P<0.001

In both men and women, greater educational attainment was associated with a lower risk of incident hypertension while greater BMI was associated with greater risk. Higher income was associated with a lower risk of incident hypertension in women. In men, smoking was associated with a greater risk of incident hypertension, while health insurance was associated with lower risks.

A competing risk regression was used to examine the incidence of hypertension while accounting for differences in mortality among race/nativity groups (Table 3). This analysis produced sub-distribution hazard ratios very similar to the hazard ratios obtained from the Cox regression. In the unadjusted model (Model 1), US-born and foreign-born Hispanic men who migrated ages 20-39 had a greater risk of incident hypertension compared to US-born White men with HRs of 1.57 (95% CI=1.25-1.98, p<0.001) and 1.45 (95% CI=1.13-1.86, p=0.004) respectively. US-born Hispanic men remained at greater risk than US-born Whites even after adjusting for socioeconomic status, health insurance, and behavioral factors (Model 4). Among women, US-born Hispanic (HR=1.54, CI=1.27-1.88, p<0.001) and foreign-born Hispanic women that migrated ages ≥ 40 years (HR=1.71, CI=1.25-2.35, p=0.001) had significantly greater risk of incident hypertension compared to US-born White women. Among US-born Hispanic Women, this increased risk no longer persisted even after adjusting for socioeconomic and health insurance (Model 3).

**Table 3.**
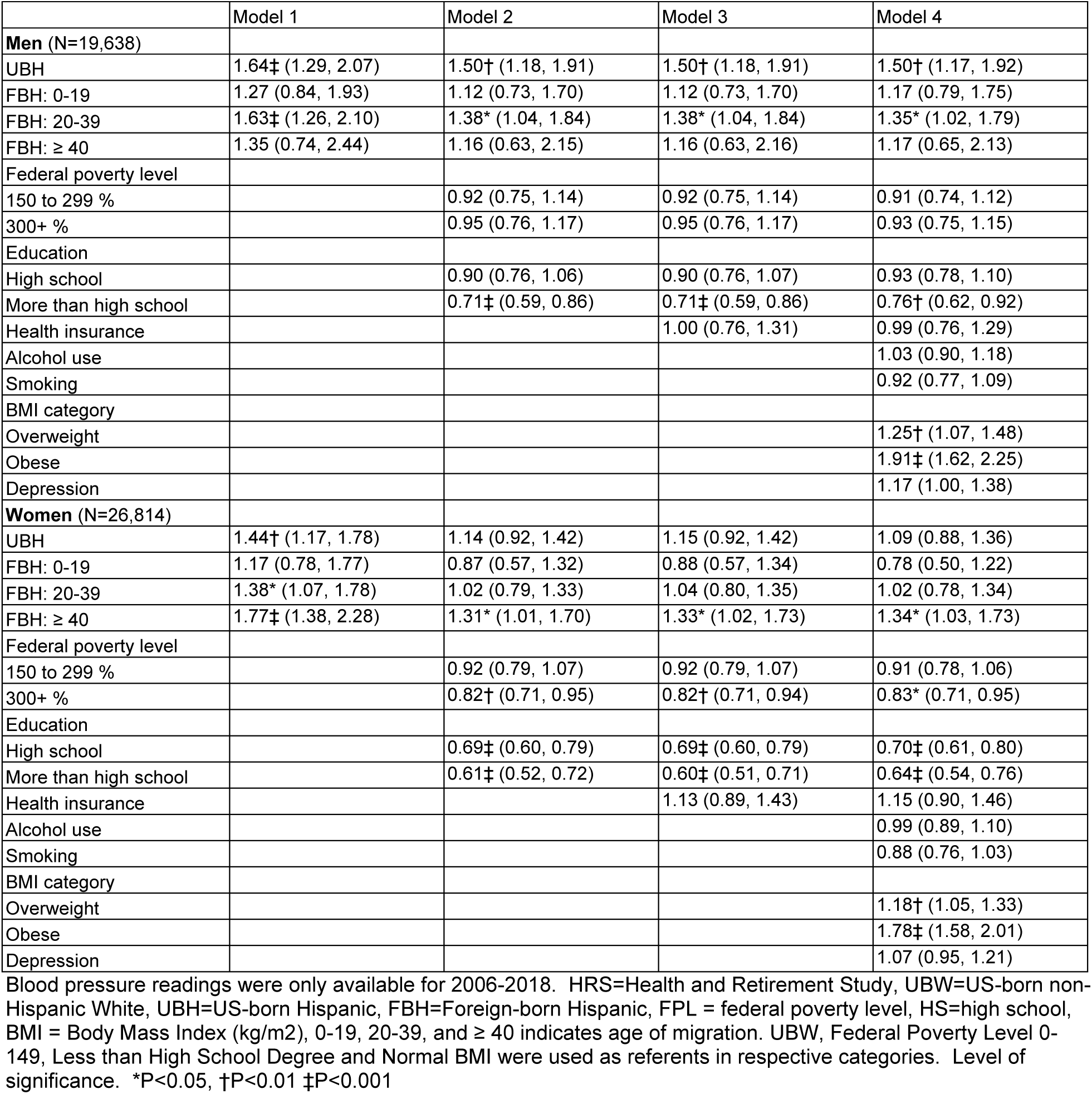
Competing risk regression for self-reported hypertension incidence. Resulted reported in sub-distribution hazard ratios. HRS 2002-2018.

|  | Model 1 | Model 2 | Model 3 | Model 4 |
| --- | --- | --- | --- | --- |
| **Men (N=19,638)** |  |  |  |  |
| UBH | 1.64‡ (1.29, 2.07) | 1.50† (1.18, 1.91) | 1.50† (1.18, 1.91) | 1.50† (1.17, 1.92) |
| FBH: 0-19 | 1.27 (0.84, 1.93) | 1.12 (0.73, 1.70) | 1.12 (0.73, 1.70) | 1.17 (0.79, 1.75) |
| FBH: 20-39 | 1.63‡ (1.26, 2.10) | 1.38* (1.04, 1.84) | 1.38* (1.04, 1.84) | 1.35* (1.02, 1.79) |
| FBH: ≥ 40 | 1.35 (0.74, 2.44) | 1.16 (0.63, 2.15) | 1.16 (0.63, 2.16) | 1.17 (0.65, 2.13) |
| Federal poverty level |  |  |  |  |
| 150 to 299 % |  | 0.92 (0.75, 1.14) | 0.92 (0.75, 1.14) | 0.91 (0.74, 1.12) |
| 300+ % |  | 0.95 (0.76, 1.17) | 0.95 (0.76, 1.17) | 0.93 (0.75, 1.15) |
| Education |  |  |  |  |
| High school |  | 0.90 (0.76, 1.06) | 0.90 (0.76, 1.07) | 0.93 (0.78, 1.10) |
| More than high school |  | 0.71‡ (0.59, 0.86) | 0.71‡ (0.59, 0.86) | 0.76† (0.62, 0.92) |
| Health insurance |  |  | 1.00 (0.76, 1.31) | 0.99 (0.76, 1.29) |
| Alcohol use |  |  |  | 1.03 (0.90, 1.18) |
| Smoking |  |  |  | 0.92 (0.77, 1.09) |
| BMI category |  |  |  |  |
| Overweight |  |  |  | 1.25† (1.07, 1.48) |
| Obese |  |  |  | 1.91‡ (1.62, 2.25) |
| Depression |  |  |  | 1.17 (1.00, 1.38) |
| **Women (N=26,814)** |  |  |  |  |
| UBH | 1.44† (1.17, 1.78) | 1.14 (0.92, 1.42) | 1.15 (0.92, 1.42) | 1.09 (0.88, 1.36) |
| FBH: 0-19 | 1.17 (0.78, 1.77) | 0.87 (0.57, 1.32) | 0.88 (0.57, 1.34) | 0.78 (0.50, 1.22) |
| FBH: 20-39 | 1.38* (1.07, 1.78) | 1.02 (0.79, 1.33) | 1.04 (0.80, 1.35) | 1.02 (0.78, 1.34) |
| FBH: ≥ 40 | 1.77‡ (1.38, 2.28) | 1.31* (1.01, 1.70) | 1.33* (1.02, 1.73) | 1.34* (1.03, 1.73) |
| Federal poverty level |  |  |  |  |
| 150 to 299 % |  | 0.92 (0.79, 1.07) | 0.92 (0.79, 1.07) | 0.91 (0.78, 1.06) |
| 300+ % |  | 0.82† (0.71, 0.95) | 0.82† (0.71, 0.94) | 0.83* (0.71, 0.95) |
| Education |  |  |  |  |
| High school |  | 0.69‡ (0.60, 0.79) | 0.69‡ (0.60, 0.79) | 0.70‡ (0.61, 0.80) |
| More than high school |  | 0.61‡ (0.52, 0.72) | 0.60‡ (0.51, 0.71) | 0.64‡ (0.54, 0.76) |
| Health insurance |  |  | 1.13 (0.89, 1.43) | 1.15 (0.90, 1.46) |
| Alcohol use |  |  |  | 0.99 (0.89, 1.10) |
| Smoking |  |  |  | 0.88 (0.76, 1.03) |
| BMI category |  |  |  |  |
| Overweight |  |  |  | 1.18† (1.05, 1.33) |
| Obese |  |  |  | 1.78‡ (1.58, 2.01) |
| Depression |  |  |  | 1.07 (0.95, 1.21) |

Blood pressure readings were only available for 2006-2018. HRS=Health and Retirement Study, UBW=US-born non-Hispanic White, UBH=US-born Hispanic, FBH=Foreign-born Hispanic, FPL = federal poverty level, HS=high school, BMI = Body Mass Index (kg/m<sup>2</sup>), 0-19, 20-39, and ≥ 40 indicates age of migration. UBW, Federal Poverty Level 0-149, Less than High School Degree and Normal BMI were used as referents in respective categories. Level of significance. \*P<0.05, †P<0.01 ‡P<0.001

### Blood pressure measurements

An analysis of blood pressure measurements was conducted on all individuals with blood pressure measurements regardless of hypertensive status. Among men, foreign-born Hispanics migrating before age 20 had a mean systolic blood pressure of 8.42 mmHg (95% CI=2.64, 14.21, p=0.005) greater than US-born White men at age 50 but had a similar change in blood pressure over time in the age adjusted model (Model 1, Table 4). This greater systolic blood pressure was no longer significantly different than US-born Whites after adjusting for income and education (Model 2). Foreign-born Hispanic women that migrated ages 0-19 and US-born Hispanic women had greater mean diastolic blood pressures of 3.75 mmHg (95% CI=0.02, 7.49, p=0.049) and 8.87 mmHg (95% CI=3.21, 14.54, p=0.003) at age 50, compared to US-born White women. This greater systolic blood pressure compared to US-born White women was no longer significantly greater in foreign-born Hispanic women migrating ages 0-19 after adjusting for socioeconomic status (Model 2) but remained greater in US-born Hispanic women even after adjusting for all factors (Model 4). Foreign-born Hispanic women that migrated at ages ≥ 40 years had a mean systolic blood pressure similar to US-born White women in the age adjusted model, however had a mean systolic blood pressure of 7.27 mmHg (95% CI=-12.98, −1.56, p=0.013) lower than US-born White women at age 50 after adjusting for socioeconomic status and remained statistically lower even after adjusting for health insurance and behavioral factors. Although foreign-born Hispanic women that migrated ages 20-39 and ≥ 40 had similar systolic blood pressure at age 50 in the age adjusted model (Model 1), they had significantly greater increases in systolic blood pressure compared to US-born White women with a mean increases 0.66 mmHg (95% CI=0.05, 1.27, p=0.035) and 0.80 mmHg (95% CI=0.16, 1.44, p=0.015) each year as they aged in the age adjusted model, as indicated by the age and race/nativity group interaction (Table 4, Model 1). This significantly greater trajectory in systolic blood pressure among foreign-born Hispanic women migrating ages ≥ 40 years was no longer significant after adjusting for socioeconomic status but again significant after adjusting for health insurance, blood pressure medication, and behavioral factors. Among foreign-born Hispanic women migrating ages 20-39 years the trajectory remained significant even after adjusting for socioeconomic status, health insurance, blood pressure medication, and behavioral factors (Model 4). Among Hispanic women migrating ages ≥ 40, this greater trajectory became more pronounced after adjusting for these risk factors.

**Table 4.**
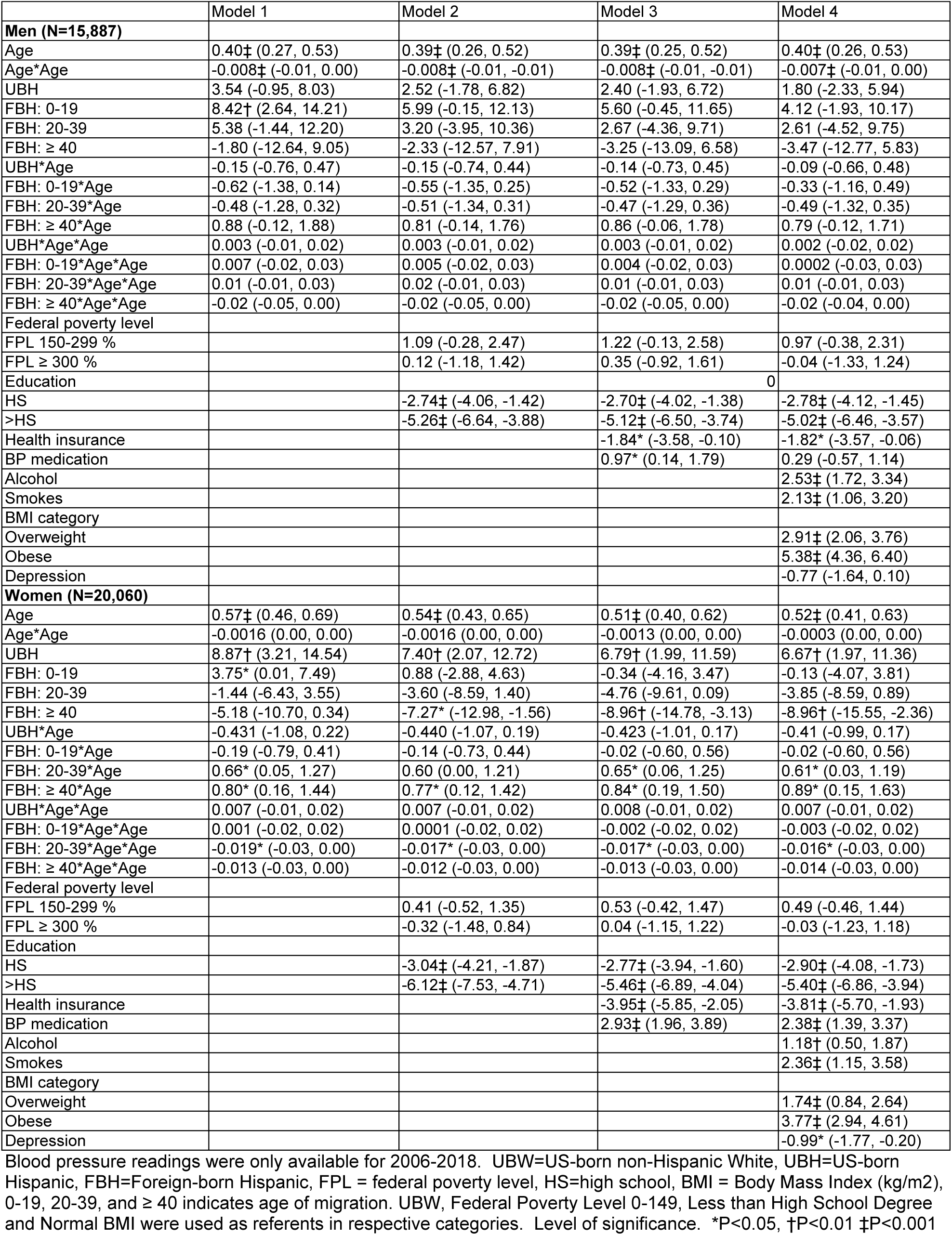
Mixed effects regression of systolic blood pressure. HRS 2006-2018.

In contrast to systolic blood pressure, diastolic pressure decreased with age. Among men, there were no differences in diastolic blood pressure among race/nativity groups (Table 5). Diastolic blood pressure of foreign-born Hispanic women migrating ages 20-39 decreased at a lower rate as they aged with an average of 0.34 mmHg (95% CI=0.03, 0.66, p=0.035) per year greater compared to US-born White women. This slower decrease with age remained after adjusting for socioeconomic status, health insurance, blood pressure medication, and behavioral factors (Model 4). Foreign-born Hispanic Women who migrated ages 20-39 had diastolic blood pressure in the age adjusted model of 2.82mmHg (95% CI=-5.64, −0.01, p=0.049) lower than US-born White women at age 50. Similarly, foreign-born Hispanic women migrating ages ≥ 40 had diastolic pressure of 3.04 mmHg (95% CI=-5.83, −0.24, p=0.034) lower than US-born White women at age 50 after adjusting for socioeconomic status. These lower diastolic blood pressures remained significant even after adjusting for health insurance, blood pressure medication, and behavioral risk factors. At age 50, US-born Hispanic women had an average diastolic blood pressure of 4.25 mmHg (95% CI=1.47, 7.03, p=0.003) greater than US-born White women in the age adjusted model (Model 1) and remained greater even after adjusting for socioeconomic, health insurance, blood pressure medication, and behavioral factors (Model 4). Greater educational attainment and health insurance were significantly associated with lower systolic and diastolic blood pressures while alcohol use and greater BMI were associated with greater blood pressures in both men and women. Smoking was associated with greater diastolic blood pressure in both men and women, and greater systolic blood pressure in women, however, was associated with lower systolic blood pressure in men. Blood pressure medication was associated with higher systolic blood pressure in women and lower diastolic blood pressure in men. This association of higher systolic blood pressure and blood pressure medication in women is likely because those with higher systolic blood pressure are selected into treatment at a higher rate than those with lower systolic blood pressure.

**Table 5.**
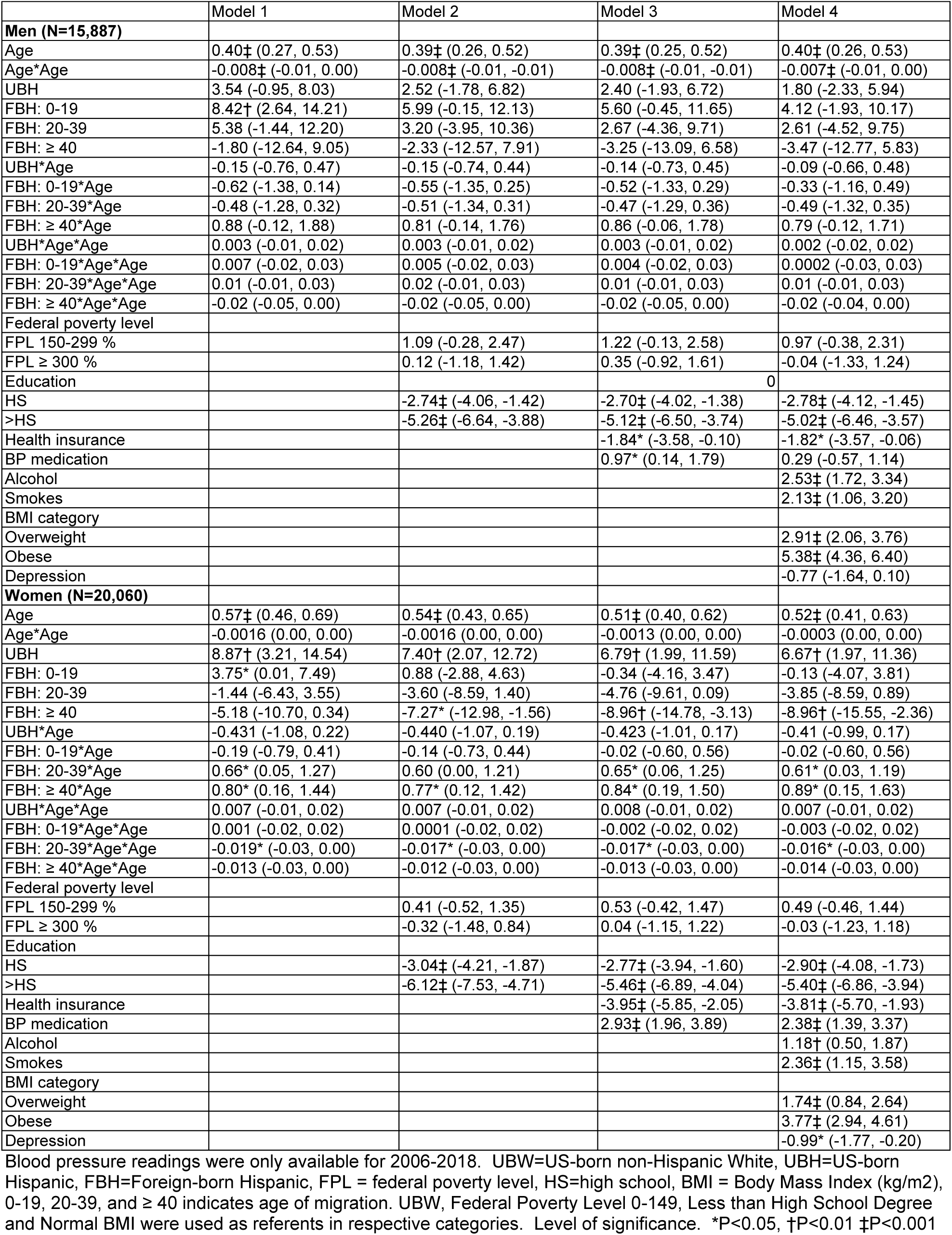
Mixed effects regression of diastolic blood pressure. HRS: 2006-2018.

**Figure 1.**
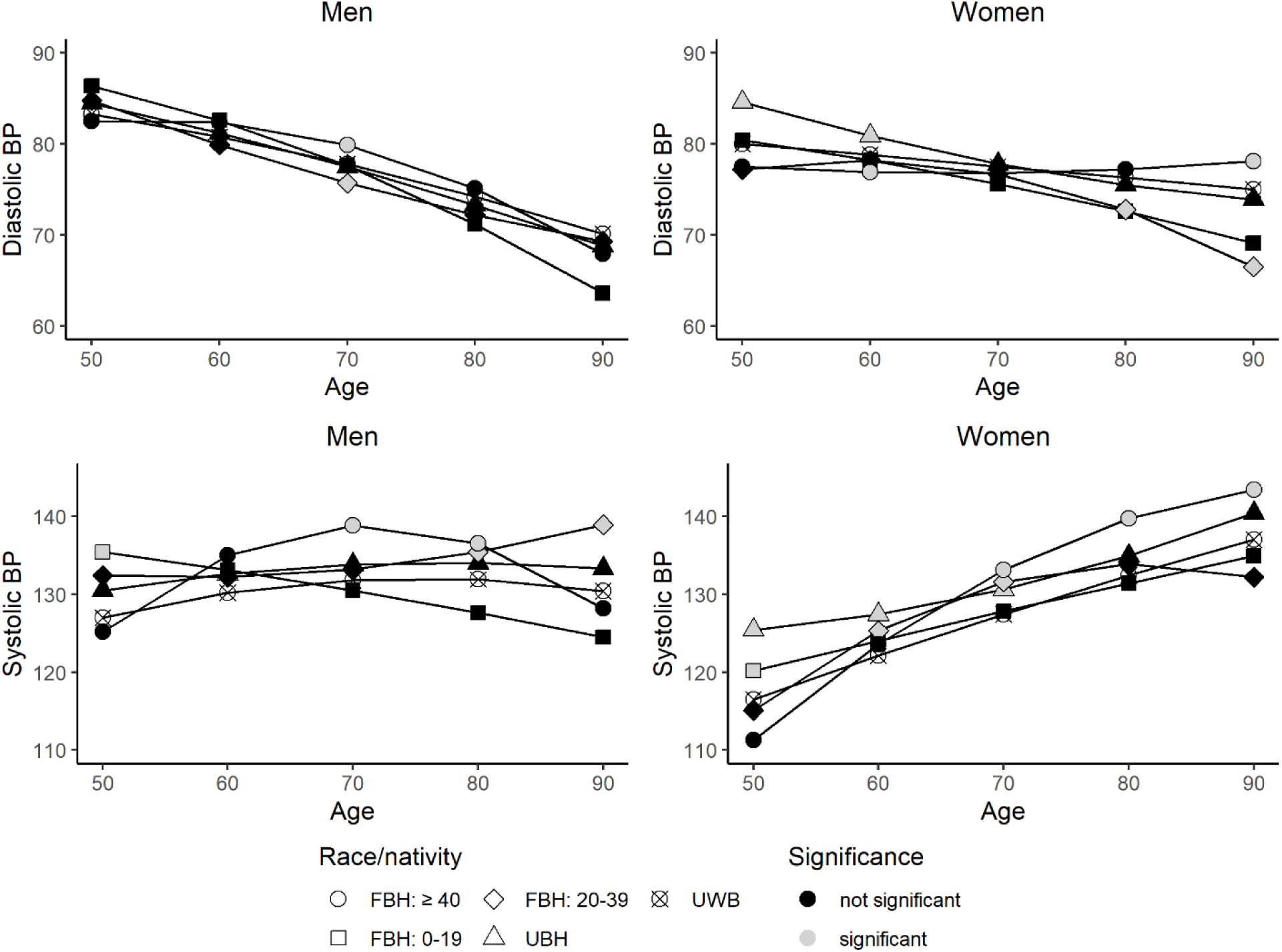
Predicted trajectories of systolic and diastolic blood pressure by sex from age-adjusted regression modeling. UBW=US-born non-Hispanic White, UBH=US-born Hispanic, FBH=Foreign-born Hispanic, 0-19, 20-39, and ≥ 40 indicates age of migration. UWB was used as the referent.

The results from Cox regression of hypertension incidence and mixed effects regressions of blood pressure indicate that socioeconomic factors explained a greater portion of health disparities of increased incidence of self-reported hypertension as well as higher blood pressures between Hispanics and US-born Whites compared to other risk factors. This greater explanatory power of socioeconomic factors was indicated in the magnitude of reduction in regression coefficients from Model 1, which included no risk factors, compared to Model 2, which included educational attainment and household income. Although the addition of health insurance in Model 3, and behavioral risk factors in Model 4 further explained disparities, reductions in coefficients when compared to Model 2 were generally smaller. Similarly, in cases where there was a protective effect such as lower blood pressure at age 50 compared to US-born Whites, the addition of socioeconomic controls further increased advantages among foreign-born Hispanics relative to US-born Whites.

## Discussion

The Hispanic paradox of lower risk of incidence of hypertension was not observed among older foreign-born Hispanic men or women regardless of age of migration. Among foreign-born Hispanic men, those who migrated between ages 20-39 were at greater risk of incident hypertension while those migrating later had similar incidence compared to US-born White men. Compared to US-born White women, foreign-born Hispanic women migrating at ages 0-19 and 20-39 had similar incidence of hypertension while those migrating ≥ age 40 had a greater incidence. These greater risks were no longer present after adjusting for income and education. At age 50, foreign-born Hispanic women that migrated ages 20-39 and ≥ 40 had similar systolic blood pressure compared to US-born White women in the age adjusted model, however, had a significantly greater increase in systolic blood pressure as they aged. These significantly greater trajectories in systolic blood pressure remained even after adjusting for socioeconomic, health insurance, and behavioral risk factors. There was also no evidence of a Hispanic paradox in hypertension risk among US-born Hispanics. Both US-born Hispanic men and women had a greater risk of incident hypertension compared to US-born Whites. These increased risks persisted even after adjusting for socioeconomic, health insurance, and behavioral risk factors. US-born Hispanic women also had greater systolic and diastolic blood pressure at age 50 compared to US-born White women even after adjusting for socioeconomic, health insurance, and behavioral risk factors, however, had similar trajectories as they aged.

Recently published research has indicated a greater prevalence of hypertension among elderly foreign-born Hispanics with less than 10 years of residency compared to US-born Whites. ^31^ Although the cited study did not examine the risk of hypertension by age of migration, the results suggested that those migrating later were at greater risk compared to US-born Whites. In contrast, the current study examined incidence rather than prevalence and only found an increased risk of incident hypertension among Hispanic women who migrated at age 40 and older. This absence of greater risk among men migrating at ≥ age 40 could be attributed to the use of self-reported hypertension in the current study, where the risk of incident hypertension among foreign-born men who migrated ≥ age 40 may be lower due to greater levels of undiagnosed hypertension. Although our smaller sample size prevented an analysis of undiagnosed hypertension by age of migration, an examination by sex indicated that foreign-born Hispanic men were at greater risk of undiagnosed hypertension compared to US-born Whites, whereas foreign-born Hispanic women were not.

Findings of greater risk of hypertension among women might be explained by diminished health selection of those that migrate later in life for family reunification purposes rather than employment. Stress associated with migration later in life may also contribute to poorer outcomes. Those who migrate later in life have a greater risk of depression that may stem from the loss of familiar social networks, language barriers, and social isolation. ^17–21^ To determine the possible role of depression in the risk of hypertension, an indicator of depression was included in our analysis based on an 8-item short version of the Center for Epidemiologic Studies-Depression (CESD) scale that was collected in the HRS. However, depression was not statistically significant and had very little effect on the regression coefficients of men or women migrating ≥ age 40 for either self-reported hypertension incidence or blood pressure measurements. This remained true even when the CESD scale was included as a continuous covariate. These findings suggest that diminished health selection among those migrating later in life may play a bigger role in the greater risk of hypertension among foreign-born Hispanic women found in the current study.

Socioeconomic status better explained the greater risk of incident hypertension and higher blood pressure among Hispanics relative to US-born Whites than behavioral factors in the current analysis. This finding mirrors those observed in a previous analysis of national, cross-sectional data (the National and suggests that addressing socioeconomic disparities between Hispanics relative to non-Hispanic Whites is an important aspect of addressing the health disparities in hypertension. ^31^ In addition to greater hypertensive risk among older Hispanics found in this study, Hispanics generally have lower levels of hypertension awareness, treatment, and control of hypertension compared to US-born Whites. ^46,47^ These disparities in hypertension control have been attributed to lower socioeconomic status and poorer access to medical care. Hispanic immigrants, especially those that migrate later may be particularly vulnerable due to relatively lower socioeconomic status and lower levels of health insurance coverage. ^48^ Policies that improve access to quality care among Hispanics could be used to address these disparities.

The analysis had the following limitations. These limitations included the use of self-reported hypertension and the potential for survivorship bias. The analysis of the incidence of hypertension relied on self-reported hypertension which may underestimate hypertension incidence among Hispanics. Although hypertension incidence based on blood pressure measurements would have provided better estimates this would have substantially reduced the sample size since these measurements are only available for 2006-2018 of the HRS and because these measurements are taken every 4 years (the core questionnaire is administered every 2 years). To address this limitation, we also examined mean blood pressure among race/nativity groups using mixed effects regression. While the analysis of hypertension incidence only included those without hypertension at baseline, the mixed effects regression included all those who received a blood pressure measurement. These results found that foreign-born Hispanic women migrating ≥ age 40 had a greater increase in mean systolic blood pressure as they aged compared to US-born White women despite having a similar mean measurement at age 50. Therefore, the findings observed in the self-reported hypertension incidence are supported by those observed in blood pressure measurements.

Although death can be treated as a censoring event in Cox regression, the analysis assumes censored data is noninformative (independent from the outcome being observed) and therefore may be biased by differences in survivorship. This assumption of independence between death and hypertension seems unlikely to be valid because hypertension increases the risk of death. To address this limitation, a competing risk regression of hypertension with death and loss to follow-up as competing risks was included in the analysis. Competing causes of death were not included in the analysis since this information it not provided in the publicly available HRS data. The competing risk regression was performed using sample weights but without standard error adjustments due to the complex sample design that is not supported for the competing risk analysis. Despite the absence of standard error adjustments in competing risk regression, these estimates produced statistically significant results with similar hazard ratios for US and foreign-born Hispanics compared to Cox regression. While this provides some evidence that our results are not driven by differences in survival, we cannot completely rule out the possibility.

## Conclusion

To our knowledge, this is the first study to use longitudinal data to examine the incidence of hypertension and blood pressure measurements among foreign-born Hispanics by age of migration to the United States. These findings indicate that the well-established Hispanic Paradox does not apply to hypertension risk among immigrants migrating later in life. Foreign-born Hispanic women who migrate ≥ age 40 are at increased risk of incident hypertension and a greater increase in systolic blood pressure as they age compared to US-born White women.

The results suggest that Hispanic women migrating later in life may be less health-selected than those migrating earlier. The results identify increased risk among a group of Hispanic immigrants that may be particularly vulnerable due to lower socioeconomic status, lower access to care, and limited eligibility for public benefits. These findings could inform medical providers of a population at greater risk of developing hypertension and policy aimed at addressing health disparities.

## Data Availability

The data was drawn from the Health and Retirement Study

https://hrs.isr.umich.edu/

